# Viral clearance as a surrogate of clinical efficacy for COVID-19 therapies in outpatients: A systematic review and meta-analysis

**DOI:** 10.1101/2023.06.18.23291566

**Authors:** Karen M Elias, Shanchita R Khan, Eva Stadler, Timothy E Schlub, Deborah Cromer, Mark N Polizzotto, Stephen J Kent, Tari Turner, Miles P Davenport, David S Khoury

## Abstract

**Background:** Surrogates of antiviral efficacy are needed for COVID-19. We investigated the relationship between the virological effect of treatment and clinical efficacy as measured by progression to severe disease in unvaccinated outpatients treated for mild to moderate COVID-19.

**Methods:** We searched PubMed, Scopus and medRxiv from inception to 27^th^ September 2022, for randomised controlled trials (RCTs) which tested potential treatments for COVID-19 in non-hospitalized patients. We included studies that reported both clinical and virological outcomes. Clinical outcomes were the rate of disease progression (generally hospitalization or death within 28 days of commencing treatment) and virological outcomes were viral load (viral RNA copies in upper respiratory tract swabs) within the first 7 days of treatment. Studies were excluded if they did not report on the outcome of a primary randomised controlled trial, or if results were reported in a more complete form in another publication. Risk of Bias assessment was performed using the RoB 2.0 tool. We used generalised linear models with random effects to assess the association between outcomes and account for study heterogeneity.

**Findings:** We identified 1372 unique studies of which 14 (with a total of 9257 participants) met inclusion criteria. Larger virological treatment effects at both day 3 and day 5 were associated with decreased odds of progression to hospitalisation or death in unvaccinated ambulatory subjects. The odds ratio (OR) for each extra two-fold reduction in viral load in treated compared to control subjects was 0.54 on both days 3 and 5 post treatment (day 3 95% CI 0.38 to 0.74, day 5 95%CI 0.41 to 0.72). There was no relationship between the odds of hospitalisation or death and virological treatment effect at day 7 (OR 0.91, 95%CI 0.74 to 1.13).

**Interpretation:** This review provides evidence that treatment-induced acceleration of viral clearance within the first 5 days after treatment is a surrogate of clinical efficacy to prevent hospitalisation with COVID-19. Limitations included the aggregation of studies with differing designs, and evidence of risk of bias in some virological outcomes. These findings support the use of viral clearance as an early phase clinical trial endpoint of therapeutic efficacy.

**Funding:** The authors were supported by the Australian Government Department of Health, Medical Research Future Fund, National Health and Medical Research Council and the University of New South Wales.

## Introduction

Several effective therapies have been developed for COVID-19, including both monoclonal antibodies and small molecule antivirals. These have been shown to be most effective when administered early in infection, with the aim of preventing progression to severe infection.^1,2^ However, the emergence of SARS-CoV-2 immune escape variants has led to a major loss of effectiveness of monoclonal antibody products, leading to reliance on a small number of antivirals.^3^ Accelerated development of new antivirals is urgently needed. While placebo-controlled trials based on clinical outcomes remain the gold standard, validated surrogate measures of treatment efficacy could accelerate efforts to develop and deploy effective COVID-19 therapies. Speeding up the process of finding and validating the effectiveness of novel COVID-19 therapies could reduce mortality especially among those most at risk.

Surrogate measures of treatment effect have been a mainstay of clinical drug development.^4^ In particular, during preclinical or early clinical development of a candidate antimicrobial treatment or monitoring of ongoing effectiveness against a potentially resistant pathogen it is common to employ a surrogate measure that is thought to predict therapeutic efficacy, such as pathogen clearance time.^5-8^ In the case of antiviral agents for COVID-19, many Phase II trials have compared the reduction in viral load between treated and control groups at different times after therapy, as a surrogate marker of therapeutic effect. For example, subjects treated with nirmatrelvir-ritonavir have been shown to have greater reduction in viral loads than controls at days 3 and 5 after treatment (0.55 and 0.80 log10 copies/mL lower respectively).^9^ Although the virological effect of treatment (the extent to which it reduces viral load compared to that seen in controls) has been widely used in studies of COVID-19 therapeutics, whether this is predictive of its clinical efficacy in preventing progression to severe outcomes has not been assessed. Here we address this question by aggregating the available studies that report the virological effects of treatment and the clinical efficacy of treatment in the same trial. We focus on analysis of treatments administered early in infection in outpatients as these have shown higher efficacy than therapies used later in hospitalised patients.^1,2^

## Results

### Characteristics of RCTs identified in literature search

We searched PubMed, Scopus and medRxiv for randomised controlled trials testing candidate COVID-19 therapies in outpatients, and which reported efficacy to prevent progression to severe COVID-19 as well as the effect of treatment on viral clearance (search terms are included in full in the supplementary methods). We identified a total of 1372 unique records of which we identified 14 eligible studies (Table S1, Figure 1).^2,9-21^ During screening we found only one study that assessed treatment in predominately vaccinated symptomatic outpatients and so we excluded this study and focused only on studies in primarily unvaccinated individuals (Figure 1). The eligible studies included six RCTs assessing small molecule antiviral therapies and eight assessing monoclonal antibodies. All 14 studies enrolled adult populations, with four of the studies also enrolling adolescents (12+ years) with risk factors for severe disease. Of the 14 studies, six excluded individuals with any prior confirmed infection, two excluded individuals who had been previously hospitalised for COVID-19, and six had no exclusion criteria based on prior infection. Most studies had criteria for inclusion of study participants that was based on risk-factors for severe COVID-19 (8/14). However, three studies selected only individuals with at least one existing risk factor for severe disease, two studies required either moderate disease or at least one risk factor if mild disease was recorded at enrolment, and one study only enrolled individuals with no risk factors. A risk of bias assessment was performed on all studies, for both clinical and virological outcomes separately, and these revealed mostly a low risk of bias (Figure S1). The only area of concern highlighted by the assessments was missing data for virological outcomes, with most studies having at least one timepoint with more than 5% of data missing, and varying levels of information about the causes of missing data.

**Figure 1.**
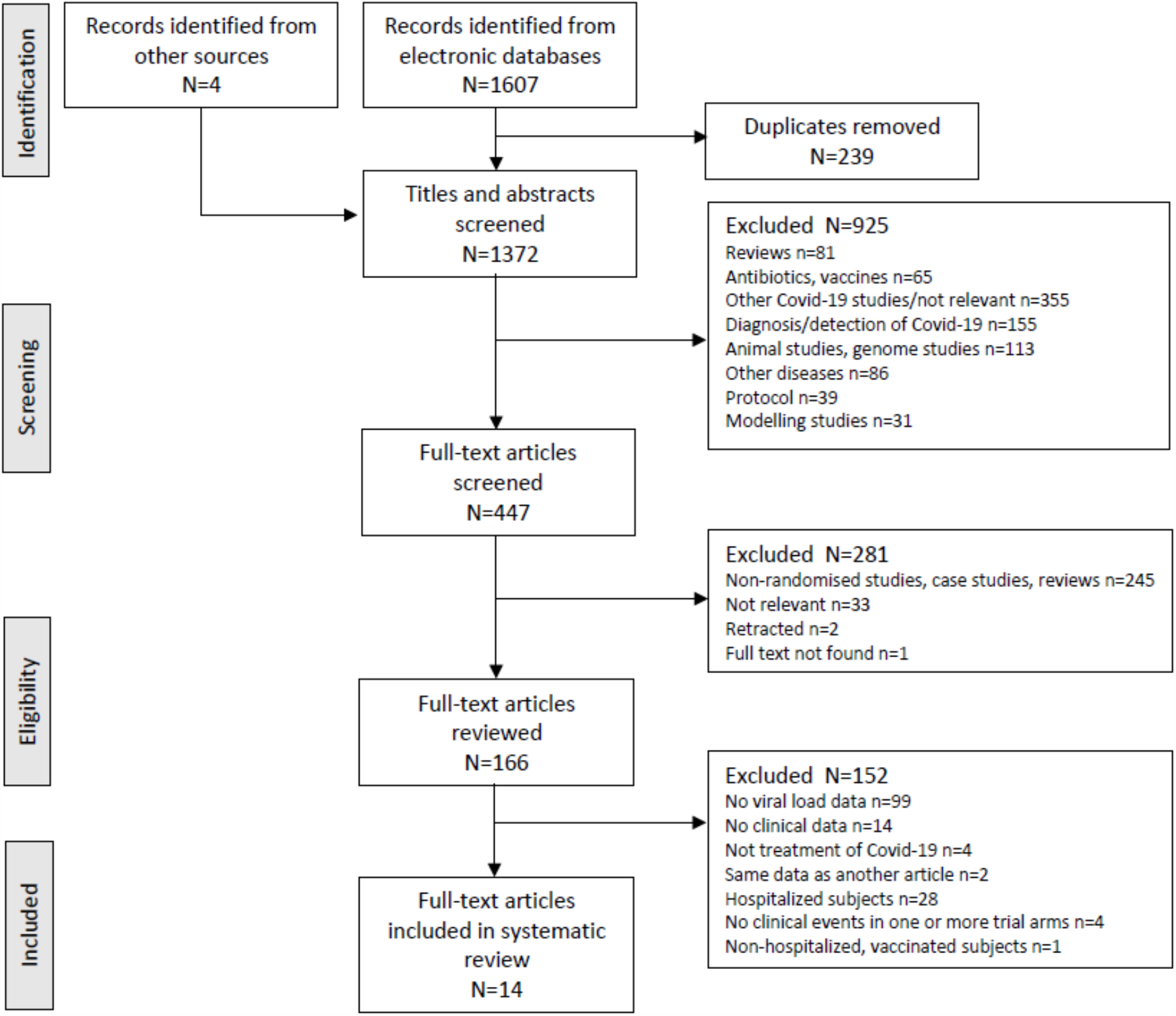
PRISMA flow diagram for study inclusion

### Clinical outcomes

The clinical endpoint reported across all studies was the relative risk of hospitalisation or death within 28 days following treatment, in the treated compared to the control group. The proportion of patients who experienced progression to hospitalisation or death in the control group ranged from 1.6% to 9.7% across all 14 studies, with a tendency for lower rates of progression in those studies with lower risk participants. Most studies were assessed as having a low risk of bias (11/14), with three studies being assessed to have some risk of bias (Figure S1A). The overall efficacy of all therapies was 58% (95% CI 48% to 65%). There was significant evidence of study heterogeneity, with the study random effect having a standard deviation (i.e., *τ*) of 0.49. Stratifying by treatment type (monoclonal antibodies vs small molecule therapies) gave similar results. The overall efficacy of monoclonal antibodies (n = 8) was 60% (95%CI 46% to 70%), similar to that of small molecule therapies (n = 6) which was 55% (95%CI 41% to 66%) (Figure 2).

**Figure 2.**
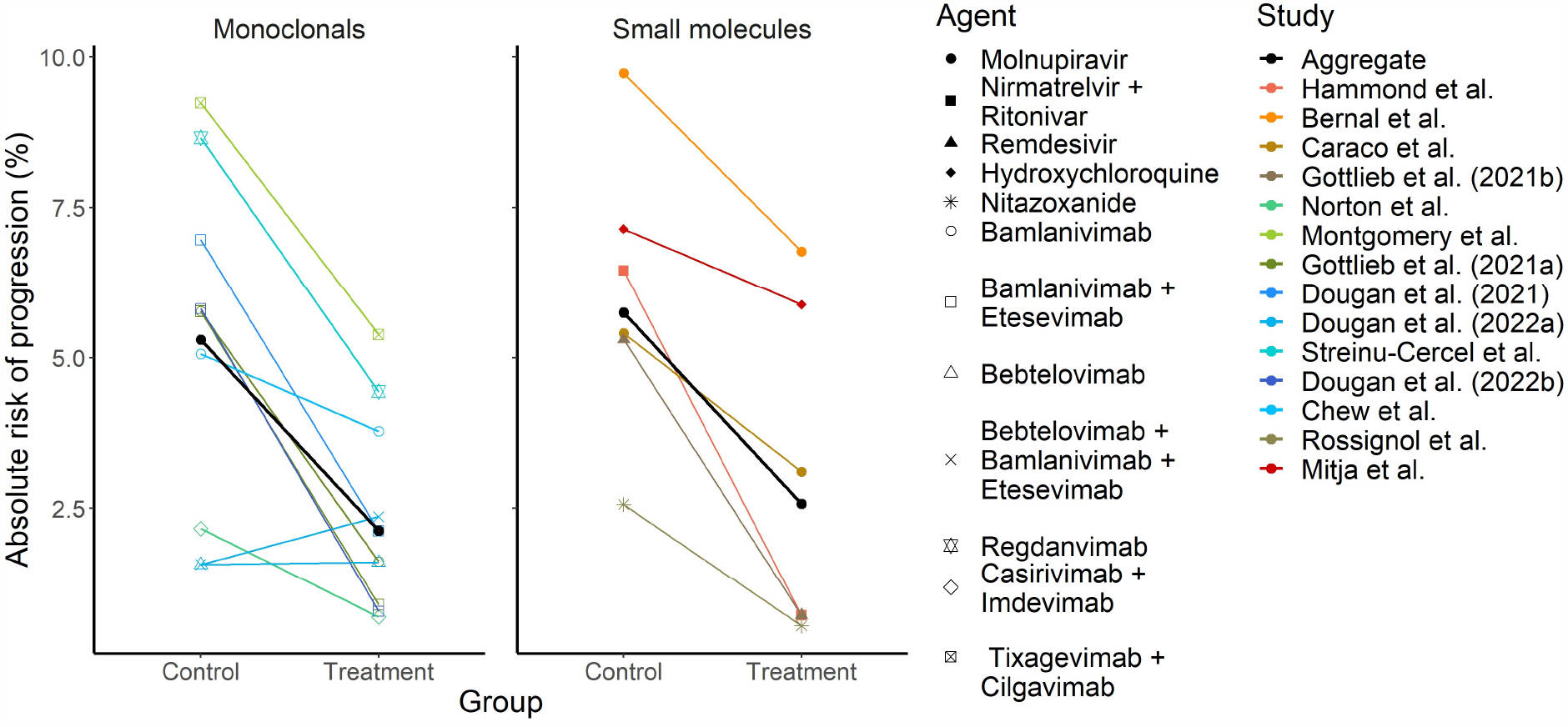
Absolute risk of progression to hospitalization or death within 28 days for treated and untreated groups for each included study, by treatment type (monoclonal antibody vs. small molecule). The absolute risk for treated and untreated groups across all included studies, stratified by treatment type is also shown (black line).

### Virological outcomes

Viral load samples were obtained from nasopharyngeal swabs in 12/14 studies, nasopharyngeal or nasal swabs in one study, and mid turbinate nasal swabs in the remaining study. Across all included studies, mean viral load decreased over time in both the treated and untreated groups (Figure 3). Relative to the day of treatment (day 1), viral load was available at a variety of different timepoints, with day 3 being the most common (n = 12/14 studies), followed by day 5 (n = 8/14) and day 7 (n = 8/14) (Figure S2). The risk of bias in virological outcomes was assessed as high in half of the studies (7/14), primarily due to missing data and reporting of the reasons for missing data (Figure S1B). All but one of the remaining studies were assessed as having some risk of bias. In studies reporting a virological outcome on day 5 the mean drop in viral load was always greater in the treated versus the control arm (n=8). In most studies reporting a virological outcome at day 3, the mean drop in viral load was greater in treated individuals compared with those in the control arm (n = 10/12 studies). We found a significant correlation between the virological treatment effect (ratio of fold-drop in viral load from baseline, in treated vs controls, see Methods Eqns. 2 and 3) measured at day 3 and day 5 (rho = 0.67, *p* = 0.046, Pearson) and day 3 and day 7 (rho = 0.92, *p* = 0.0002). However, we did not observe a significant correlation between the virological treatment effect on day 5 and day 7 (rho = 0.38, *p* = 0.46), perhaps due to limited power to detect a correlation (Figure S3). Further, the virological treatment effect tended to approach the maximum observed effect size within the first 7 days (Figure 3). That is, for studies where a virological treatment effect was seen, and where data was available for any timepoint after day 7 (n=11/14), the maximum observed virological treatment effect occurred on or before day 7 in all except one study (i.e., n=10/11). Together this indicates that the largest observed virological treatment effect tends to occur on or before day 7.

**Figure 3.**
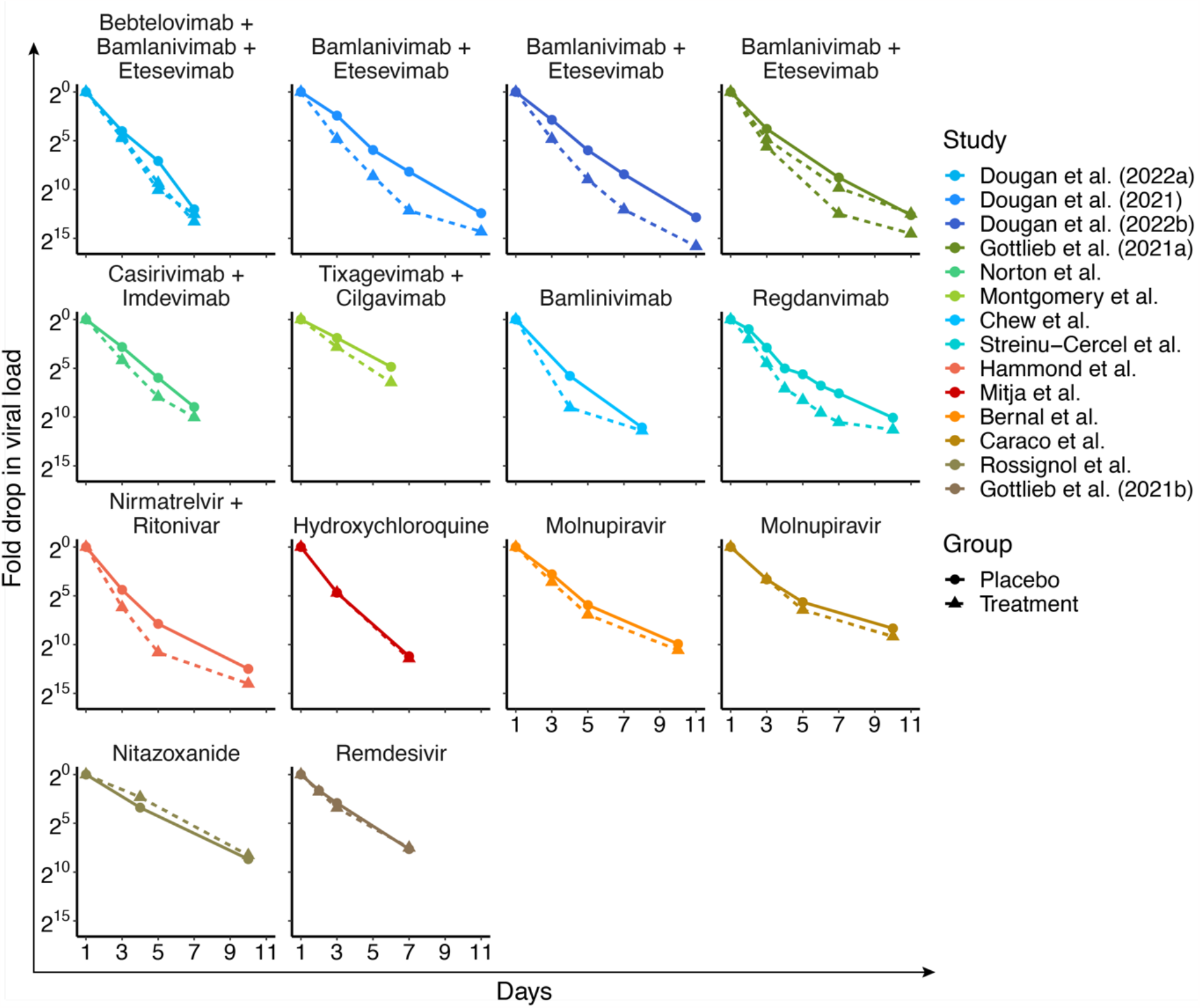
Change in viral load over time for each study in treatment (dashed line) and control (solid line) groups. Gottlieb et al (2021a) has two treatment groups, one received Bamlinivimab alone, the other received Bamlinivimab and Etesevimab, Dougan et al. (2022a) has two treatment groups, one received Bebtelovimab alone, the other received Bebtelovimab together with Bamlinivimab and Etesevimab.

### Relationship between virological treatment effect and clinical efficacy

To investigate the relationship between clinical and virological outcomes, we used a generalised linear mixed model (GLMM). We compared this relationship using virological treatment effect assessed at days 3, 5 and 7, separately. The models using data at day 3 (which included 12 studies) and at day 5 (which included 8 studies) produced broadly consistent results, with both showing a significant relationship (Table S2). Each two-fold increase in viral clearance (i.e., one Ct value by PCR) in treated individuals compared with control individuals was associated with reduced odds of disease progression (day 3: OR 0.54, 95%CI 0.38 to 0.74, p = 0.0002; day 5: OR 0.54, 95%CI 0.41 to 0.72, p < 0.0001). However, we did not find evidence that increased virological treatment effect at day 7 was associated with improved clinical outcomes (OR 0.91, 95%CI 0.74 to 1.13, p = 0.39). For each model, we converted the model estimates to an efficacy (see Methods, Eqns. 5 and 6) and visualised the relationship between virological treatment effect and clinical efficacy (Figure 4).

**Figure 4.**
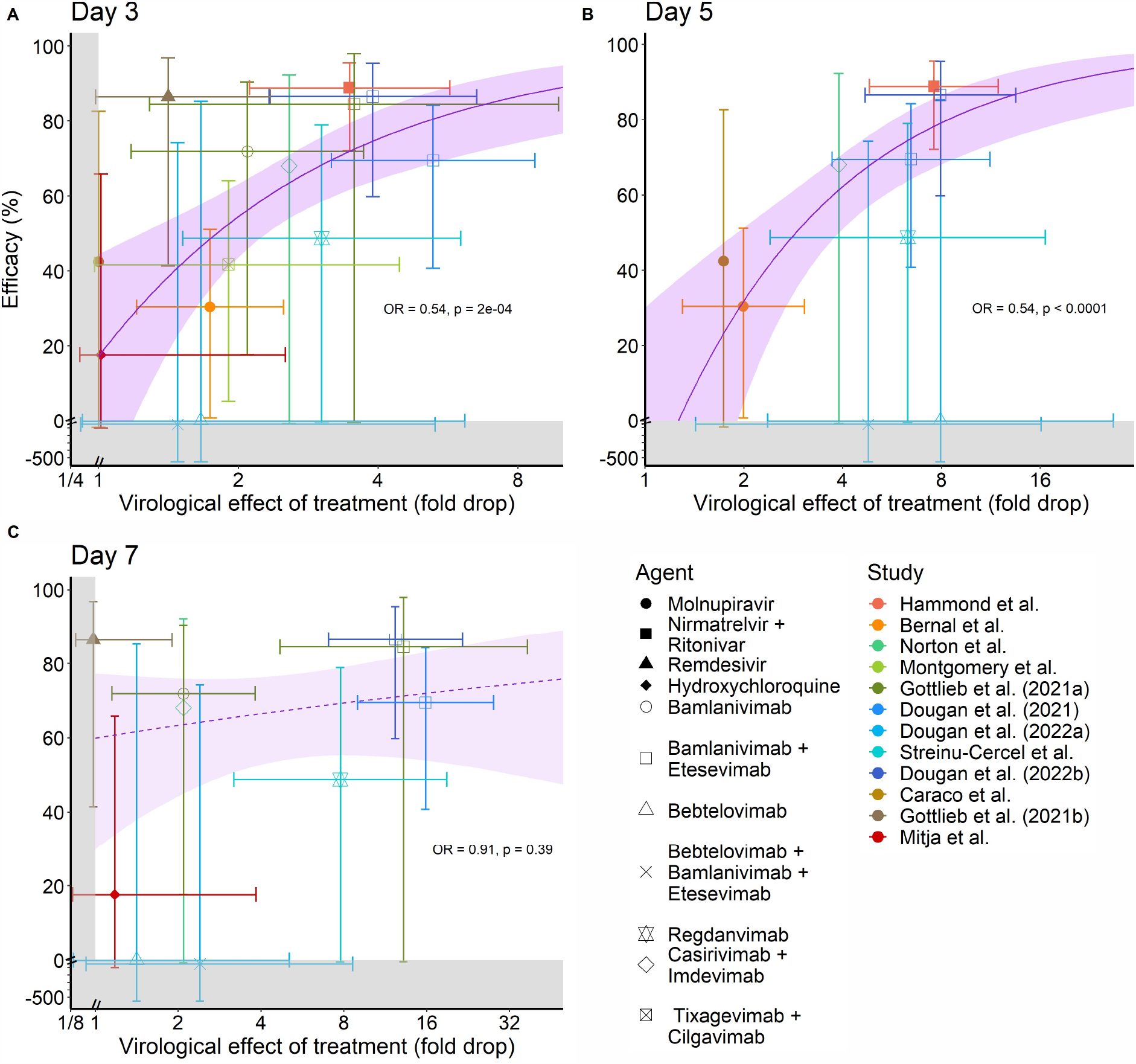
Correlation between virological effect at (A) day 3, (B) day 5, and (C) day 7 and clinical efficacy to prevent hospitalization in unvaccinated COVID-19 outpatients treated with a small molecule antiviral or monoclonal antibody. Error bars of each data point indicate 95% confidence intervals. The solid and dashed lines indicate the fitted model regression models with 95% confidence regions indicated by shading. A dashed line was used where the relationship was not found to be significant (C).

We assessed the robustness of these associations to other potential confounders such as treatment type (mAb or small molecule) and variant. These models showed no evidence of an effect from these co-variates and produced very similar results to the unadjusted model (Table S2). Further, we undertook sensitivity analysis to compare results obtained from those studies with a low to medium risk of bias, and those with a high risk of bias, and found no difference in the conclusions reached (Table S3).

To assess whether the virological treatment effect measured on day 3 or day 5 was a better predictor of clinical effect, we compared the model fits obtained using data from each of these days. To ensure consistent study inclusion in this comparison, we refitted these models using only those studies that reported data on both day 3 and 5 (n = 8). We found that the virological treatment effect at day 3 and day 5 were similarly predictive of efficacy, but with the day 5 timepoint providing a slightly better model fit (AIC 108.0 for day 5 vs. 111.8 for day 3). Finally, we also considered various composite measures of virological treatment effect, namely the maximum virological effect observed within 7 days, the average of observed virological effects up to day 6 (inclusive), and the difference in viral clearance rate to day 6 between treated and controls, to incorporate data from all clinical trials regardless of sampling days. We found that all these composite measures also showed a significant correlation between virological effect and clinical effect (Table S2, Figure S4), providing evidence that the exclusion of studies with different sampling times was not biasing our results. Together this shows that virological treatment effect over short timeframes, within five days of commencing treatment, is a useful predictor of clinical outcomes.

## Discussion

Rapid virological clearance is often thought to be a positive prognostic indicator. In COVID-19, higher viral load and poor virological control tends to correlate with more severe infection outcomes ^13^. However, despite the frequent use of virological outcomes as phase I/II clinical trial endpoints for therapies, these have rarely been rigorously shown to be correlated with clinical outcomes of relevance, such as reduced risk of hospitalisation. Here we show that for treatments administered to unvaccinated COVID-19 outpatients, the treatment effect to reduce SARS-CoV-2 RNA levels in upper respiratory tract swabs is correlated with clinical efficacy to prevent hospitalisation and death. We found a robust association between virological treatment effect observed at day 3 and 5 and clinical outcomes. Thus, virological clearance represents an important surrogate of clinical efficacy, especially in early-stage clinical trials.

Our study included a variety of potential treatments, with differing mechanisms of action. Although we show an association between virological and clinical effects, it is also plausible that some novel treatments may affect clinical outcomes without impacting virological outcomes in upper respiratory tract samples. This suggests that a lack of observed virological efficacy may not necessarily indicate a lack of clinical efficacy, and this will be especially true in host targeting therapies aimed, for example, at reducing pathogenic immune responses, which are not included in this analysis. However, our results also indicate that the relationship between virological outcomes and clinical outcomes may constitute a tool for predicting a minimum expected efficacy for a given observed virological effect (Figure S5). For example, if a novel agent were shown to induce at least an extra 2.3-fold drop in viral load by day 3 (i.e., a 1.2 increase in Ct value), this is expected to be associated with a 50% clinical protection from hospitalisation (on the lower bound of 95% confidence interval of predicted efficacy). Since virological outcomes can be assessed in low-risk populations, conceivably even in a placebo-controlled trial, this may represent a means of predicting efficacy for at risk individuals without needing to run a placebo-controlled trial in the most vulnerable populations.

We observed that virological outcomes at day 5 were slightly better predictors of clinical outcomes than those at day 3. However, this could only be examined in 8/14 studies, and the differences were not dramatic. Since virological treatment effect measured at day 3 and 5 were correlated, it is likely that the most robust metric of virological effect will be some composite measure that uses information across multiple days. However, constructing and validating such a composite measure is difficult from the available data, since only a subset of studies has detailed data on viral loads over time. Thus, we suggest that in future phase I and II clinical trials aiming to provide an indication of eventual clinical effect of a treatment, virological outcomes should be measured on at least days 1 (i.e., on the day of first treatment), 3 and 5 to determine whether a virological treatment effect is observed at both time points. We did not find evidence of a relationship between virological outcomes at day 7 and clinical outcomes. This is likely due to a combination of fewer studies having data available at this timepoint, and viral loads being lower and starting to converge between treated and controls by this point in some studies (Figure 3).

An important limitation of this study is that our analysis has only considered studies of COVID-19 therapeutics in unvaccinated, naïve and (largely) immunocompetent populations, whereas these drugs are currently most commonly being used in vaccinated and / or immunosuppressed people. While this is a significant issue for our study and the use of a virological surrogate, it is also a broader issue for the field since we are at present using agents in populations for which efficacy has not been directly demonstrated. Conflicting evidence exists as to treatment efficacy in vaccinated populations. The PANORAMIC trial ^22^ studied the effects of molnupiravir in a largely (97%) vaccinated population and found no significant protection from hospitalisation or death, in contrast to the original studies that reported 30% ^17^ and 42% ^10^ efficacy in unvaccinated individuals. However, a recent observational study showed no effect of vaccination status on the efficacy of nirmatrelvir-ritonavir (although lower overall efficacy was observed than in the RCT analysed here).^23^ More studies of therapies in vaccinated/non-naïve populations are needed to test the extent to which the observed relationship between virological and clinical outcomes may be altered by vaccination.

An additional limitation is that, despite our systematic search of the literature, a risk of publication bias remains. This is because there is a higher likelihood of publication of trial results that show a significant treatment effect. Further, there were methodological differences between the trials, such as timing of commencement of treatment (days post-symptom onset), as well as timing and site of swab collection for viral load measurement (Table S1), and other potential study design differences may have been present that could not be adequately measured or controlled for in our analysis.

Importantly, we have not proven that the correlation between a virological treatment effect and improved clinical outcomes described here is the causal mechanism of protection of these therapies. However, this seems highly plausible given links between higher viral load and disease severity ^13^. In fact, our analysis combines data from multiple agents with different mechanisms of action, e.g., monoclonal antibodies (8/14 studies) and a range of small molecules (6/14). It is possible that different mechanisms of action may show different relationships, and although we did not see an impact of treatment type on these relationships (Table S2), our study had limited power to detect these different relationships across different agents if they were present.

Together, this work provides an evidence base for the use of virological outcomes as surrogates of clinical outcomes, such as hospitalisation, for COVID-19. Viral outcomes represent an important potential predictor of clinical outcomes during early-stage clinical trials of novel therapeutics for viral infections of both COVID-19, and potentially other viruses of the upper respiratory tract, and may accelerate discovery and approval of antiviral treatments.

## Methods

### Search strategy and selection criteria

Our systematic search of the literature followed the Preferred Reporting Items for Systematic reviews and Meta-Analysis (PRISMA) statement for study design (see supplementary file, PRISMA checklist).^24,25^ We searched PubMed, Scopus and medRxiv for randomized placebo-controlled trials of various treatments for COVID-19 in non-hospitalised patients from inception until September 27^th^, 2022 (see Supplementary Methods for full search strategy). Articles were screened to determine eligibility for final inclusion (SRK), using inclusion and exclusion criteria outlined in the supplementary material. Publications that were reviews or protocols, animal, or in-vitro studies, observational or case studies, studies on vaccines, host-directed therapies or antibiotics were excluded. We included all identified RCTs testing virus directed COVID-19 therapies in non-hospitalized and unvaccinated individuals, and which reported both viral load data for at least one timepoint less than day 7, and rates of progression to either hospitalisation or death (Table S1). We identified 14 studies for inclusion with a total of 9257 participants. From these studies we extracted all data on viral load at baseline and after treatment at all available timepoints up to 14 days, for treatment and control arms of the studies, as well as number of individuals in each arm and the number of outcomes (i.e., progression of disease as defined by the study and summarised in Table S1). Other study data that was collected included, inclusions/exclusion criteria for participants in each of the RCTs, treatment administered (including dosing and timing information) and viral load quantification method (Table S1). Data extraction from the selected studies was performed independently by two researchers (KME and SRK). Where data was not provided in tables or text, it was extracted from figures using WebPlotDigitizer (Version 4.6), discrepancies were resolved through discussion and consensus amongst data extractors. The revised tool for risk of bias in randomised trials (RoB 2.0) was used to assess the studies.^26,27^ The risk of bias assessment of the included studies was carried out by KME and SRK independently and any disagreements were resolved through discussion. A systematic review protocol was not pre-registered for this review.

### Clinical effect of treatment

Disease progression was generally classified as hospitalisation due to COVID-19 or death from any cause for a period of 28 days after the commencement of treatment, with some differences between studies i.e., using 28 or 29 days of follow-up and one study counting as progression Emergency Department (ED) presentation in addition to hospitalisation in their clinical efficacy (Table S1). Clinical outcome data was available as count data showing the number of subjects who progressed to hospitalization or death for each trial arm. This count data was used to calculate the clinical effect of treatment for each study as an Odds Ratio (OR). OR were converted to a relative risk (RR) and efficacy (100 × (1 − *RR*)) for visualisation of the data and analysis. If a trial contained treatment with more than one monoclonal antibody combination, we considered different combinations separately (e.g., Bamlanivimab vs. Bamlinivimab and Etesevimab). However, if a trial contained multiple doses of the same treatment (i.e., same drug, or same monoclonal combination), then results of the same treatment at different doses were pooled. Clinical outcome data was aggregated using a mixed effects logistic regression model (using the glmer function from the lme4 package in R version 4.2.1) to model disease progression as a binary outcome and grouped by small molecules and monoclonal antibodies. Random effects were used to account for study heterogeneity and quantified by the standard deviation of these effects (*τ*). The model used is given by:

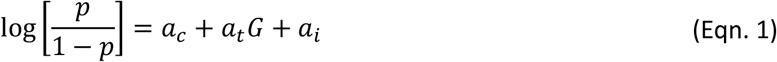

where *p* is the proportion of events in the group (i.e. *n*_*t*_/*N*_*t*_ for treated group and *n*_*c*_/*N*_*c*_ for control group where *n* is the number of events, and *N* is the number at risk), *a*_*c*_ is the log odds of disease progression in the controls across all studies, *a*_*t*_ is the log(OR) of disease progression in the treated compared to controls across all studies, G is an index variable taking the value 0 for control groups and 1 for treatment groups, and *a*_*i*_ are the random effects for each study, *i* (assumed normally distributed around mean 0 with standard deviation, *τ*).

The vertical error bars in Figures 4 and S4 show 95% confidence intervals for the clinical efficacy which were calculated using the Katz-log method.^28^

### Virological effect of treatment

For any study group (*G*) at day *d*, the fold-drop from baseline (day 1) was calculated as:

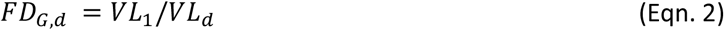

where *VL*_*d*_ is the viral load at day *d*. The virological treatment effect (associated with day *d*) for all treatment groups was defined as, the

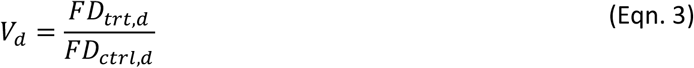

where *FD*_*trt,d*_ and *FD*_*ctrl,d*_ are the fold drop in the treated and control groups from day 1 to day *d*, respectively. This was calculated at each day (*d*) where viral load data was provided in each study. In Figure 4, the horizontal error bars are 95% confidence intervals derived from published data on the spread of the virological treatment effect (where possible). Points from studies where this was not possible are depicted without horizontal error bars. The methods used to standardise these horizontal error bars are summarized in Table S3.

### Relationship between virological treatment effect and clinical efficacy

To assess whether there was a significant relationship between virological and clinical effect of treatments in the available data, we fit three separate models. These models incorporated virological treatment effects as measured either at day 3, day 5, or day 7 as the predictor. We used a mixed effects logistic regression model (using the glmer function from the lme4 package in R version 4.2.1) to model disease progression as a binary outcome. The model included a random effect on the intercept for each study. Specifically, we fit the regression model:

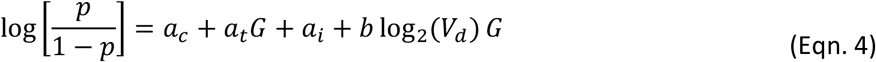

where *V*_*d*_ is the mean virological treatment effect for each treatment group at day *d*, and b is the log(OR) of each 2-fold increase in the virological effect, and other terms in Eqn. 4 are as defined in Eqn. 1.

We used the model to estimate the log(OR) for treated groups relative to the baseline log odds of progression in the untreated groups, which were then converted to (absolute) risk for control and treated groups, as well as relative risk (RR) using:

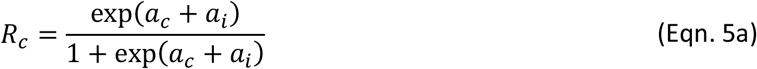

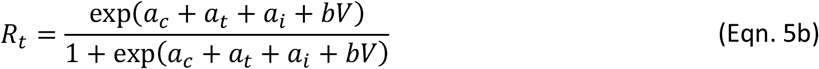

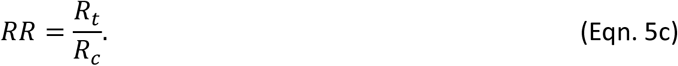

Efficacy was then calculated as:

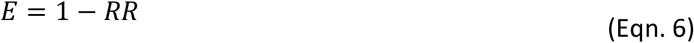

For visualisation in Figure 4, we plotted the marginal mean to show the average effect over all potential studies (i.e., assumed *a*_*i*_ = 0).

### Determining Confidence Intervals

To obtain confidence intervals for the estimated clinical efficacy for a given level of virological effect we performed parametric bootstrapping. We used the parameter estimates and covariance matrix obtained from maximum likelihood model fitting to describe a multivariate normal distribution (using the function rmvnorm from package mvtnorm), which was then sampled 10,000 times. We then calculated the efficacy from these sampled values at each level of virological effect and used percentiles of the results to estimate the lower and upper bound 95% confidence intervals.

## Supporting information

PRISMA Checklist

PRISMA Checklist (abstract)

## Data Availability

Extracted data and codes for analysis will be made publicly available on GitHub upon publication.

## Ethics statement

This work was approved under the UNSW Sydney Human Research Ethics Committee (approval HC200242).

## Data and code availability

Extracted data and codes for analysis will be made publicly available on GitHub upon publication.

## References

1. Stadler E, Chai KL, Schlub TE, et al. Determinants of passive antibody efficacy in SARS-CoV-2 infection. medRxiv 2022: 2022.03.21.22272672.

2. Montgomery H, Hobbs FDR, Padilla F, et al. Efficacy and safety of intramuscular administration of tixagevimab-cilgavimab for early outpatient treatment of COVID-19 (TACKLE): a phase 3, randomised, double-blind, placebo-controlled trial. Lancet Respir Med 2022.

3. Toussi SS, Hammond JL, Gerstenberger BS, Anderson AS. Therapeutics for COVID-19. Nature Microbiology 2023; 8(5): 771–86.

4. FDA. Surrogate endpoint resources for drug and biologic development. https://www.fda.gov/drugs/development-resources/surrogate-endpoint-resources-drug-and-biologic-development (accessed 05 June 2023).

5. Perelson AS, Guedj J. Modelling hepatitis C therapy--predicting effects of treatment. Nat Rev Gastroenterol Hepatol 2015; 12(8): 437–45.

6. Barber BE, Fernandez M, Patel HB, et al. Safety, pharmacokinetics, and antimalarial activity of the novel triaminopyrimidine ZY-19489: a first-in-human, randomised, placebo-controlled, double-blind, single ascending dose study, pilot food-effect study, and volunteer infection study. Lancet Infect Dis 2022; 22(6): 879–90.

7. Dondorp AM, Nosten F, Yi P, et al. Artemisinin Resistance in Plasmodium falciparum Malaria. New England Journal of Medicine 2009; 361(5): 455–67.

8. McCarthy JS, Yalkinoglu Ö, Odedra A, et al. Safety, pharmacokinetics, and antimalarial activity of the novel plasmodium eukaryotic translation elongation factor 2 inhibitor M5717: a first-in-human, randomised, placebo-controlled, double-blind, single ascending dose study and volunteer infection study. Lancet Infect Dis 2021; 21(12): 1713–24.

9. Hammond J, Leister-Tebbe H, Gardner A, et al. Oral Nirmatrelvir for High-Risk, Nonhospitalized Adults with Covid-19. N Engl J Med 2022; 386(15): 1397–408.

10. Caraco Y, Crofoot GE, Moncada PA, et al. Phase 2/3 Trial of Molnupiravir for Treatment of Covid-19 in Nonhospitalized Adults. NEJM Evidence 2022; 1(2): EVIDoa2100043.

11. Chew KW, Moser C, Daar ES, et al. Antiviral and clinical activity of bamlanivimab in a randomized trial of non-hospitalized adults with COVID-19. Nature Communications 2022; 13(1): 4931.

12. Dougan M, Azizad M, Chen P, et al. Bebtelovimab, alone or together with bamlanivimab and etesevimab, as a broadly neutralizing monoclonal antibody treatment for mild to moderate, ambulatory COVID-19. medRxiv 2022: 2022.03.10.22272100.

13. Dougan M, Azizad M, Mocherla B, et al. A Randomized, Placebo-Controlled Clinical Trial of Bamlanivimab and Etesevimab Together in High-Risk Ambulatory Patients With COVID-19 and Validation of the Prognostic Value of Persistently High Viral Load. Clin Infect Dis 2022; 75(1): e440–e9.

14. Dougan M, Nirula A, Azizad M, et al. Bamlanivimab plus Etesevimab in Mild or Moderate Covid-19. New England Journal of Medicine 2021; 385(15): 1382–92.

15. Gottlieb RL, Nirula A, Chen P, et al. Effect of Bamlanivimab as Monotherapy or in Combination With Etesevimab on Viral Load in Patients With Mild to Moderate COVID-19: A Randomized Clinical Trial. JAMA 2021; 325(7): 632–44.

16. Gottlieb RL, Vaca CE, Paredes R, et al. Early Remdesivir to Prevent Progression to Severe Covid-19 in Outpatients. New England Journal of Medicine 2021; 386(4): 305–15.

17. Jayk Bernal A, Gomes da Silva MM, Musungaie DB, et al. Molnupiravir for Oral Treatment of Covid-19 in Nonhospitalized Patients. New England Journal of Medicine 2021; 386(6): 509–20.

18. Mitjà O, Corbacho-Monné M, Ubals M, et al. Hydroxychloroquine for Early Treatment of Adults With Mild Coronavirus Disease 2019: A Randomized, Controlled Trial. Clin Infect Dis 2021; 73(11): e4073–e81.

19. Norton T, Ali S, Sivapalasingam S, et al. REGEN-COV Antibody Combination in Outpatients With COVID-19 – Phase 1/2 Results. medRxiv 2022: 2021.06.09.21257915.

20. Rossignol J-F, Matthew CB, Oaks JB, et al. Early treatment with nitazoxanide prevents worsening of mild and moderate COVID-19 and subsequent hospitalization. medRxiv 2021: 2021.04.19.21255441.

21. Streinu-Cercel A, Săndulescu O, Preotescu LL, et al. Efficacy and Safety of Regdanvimab (CT-P59): A Phase 2/3 Randomized, Double-Blind, Placebo-Controlled Trial in Outpatients With Mild-to-Moderate Coronavirus Disease 2019. Open Forum Infect Dis 2022; 9(4): ofac053.

22. Butler CC, Hobbs FDR, Gbinigie OA, et al. Molnupiravir plus usual care versus usual care alone as early treatment for adults with COVID-19 at increased risk of adverse outcomes (PANORAMIC): an open-label, platform-adaptive randomised controlled trial. Lancet 2022.

23. Aggarwal NR, Molina KC, Beaty LE, et al. Real-world use of nirmatrelvir-ritonavir in outpatients with COVID-19 during the era of omicron variants including BA.4 and BA.5 in Colorado, USA: a retrospective cohort study. Lancet Infect Dis 2023; 23(6): 696–705.

24. Liberati A, Altman DG, Tetzlaff J, et al. The PRISMA statement for reporting systematic reviews and meta-analyses of studies that evaluate healthcare interventions: explanation and elaboration. BMJ 2009; 339: b2700.

25. Moher D, Liberati A, Tetzlaff J, Altman DG. Preferred reporting items for systematic reviews and meta-analyses: the PRISMA statement. BMJ 2009; 339: b2535.

26. Higgins JP, Savović J, Page MJ, Elbers RG, Sterne JA. Assessing risk of bias in a randomized trial. Cochrane Handbook for Systematic Reviews of Interventions; 2019: 205–28.

27. Sterne JAC, Savović J, Page MJ, et al. RoB 2: a revised tool for assessing risk of bias in randomised trials. BMJ 2019; 366: 4898.

28. Aho K, Bowyer R. Confidence Intervals for Ratios of Proportions: Implications for Selection Ratios. Methods in Ecology and Evolution 2014; 6.

