## Supplementary Material for "Viral clearance as a surrogate of clinical efficacy for COVID-19 therapies in outpatients: A systematic review and meta-analysis"

### Supplementary methods

#### Search Strategy

A systematic search of the literature was performed using PubMed, Scopus and medRxiv from their inception to September 27th, 2022. The PICOS framework (Population, Intervention, Control, Outcomes and Study type) was adapted to define the inclusion and exclusion criteria. Search terms were:

("COVID-19" OR "SARS-CoV-2" OR "coronavirus") AND

("monoclonal antibody" OR "antiviral" OR "convalescent plasma") AND

("viral load" OR "viral clearance" OR "viral elimination")

Language was limited to English.

#### Inclusion criteria

Randomised controlled trials where the population was COVID-19 outpatients, any monoclonal antibodies, convalescent plasma, or antivirals used as an intervention, viral load data and clinical outcome data reported as outcome were included. Both peer-reviewed and pre-print studies were included. If a study had a published version available of a pre-print version found in the searches, the data was extracted from the final published version.

#### Exclusion criteria

Publications were excluded if they were:

- reviews or protocols,
- animal or in-vitro studies,
- observational or case studies,
- studies on vaccines or antibiotics or host directed therapies,
- viral load data was not reported for at least one timepoint less than 7 days after treatment, or
- where the population was hospitalised were excluded from the systematic review.

If two studies contained results on the same set of subjects, only the paper with the most data available was used. We subsequently excluded four studies where there were no clinical events in one or more trial arms, as the lack of events indicated small sample sizes, and high uncertainty in these studies. We determined that only one of the 15 remaining studies included vaccinated subjects, so we excluded this study and focused on investigating the relationship in unvaccinated patients only.

**Table S1** Data sources for clinical and virological efficacy data

| Paper (ref) | Treatment and dosing | Population description | Clinical outcome used | Viral load assessment method | Viral load assessed (days) | Virological data source | Clinical data source |
|---|---|---|---|---|---|---|---|
| Hammond et al. [1] | Nirmatrelvir + Ritonavir vs. placebo (twice daily for 5 days) | Outpatients at high risk of severe disease[a] | COVID-19 related hospitalization or death from any cause by day 28 | Nasopharyngeal or nasal swabs. Reported as adjusted mean change from baseline in $\log_{10}$ copies/mL | 1,3,5,10,14 | Figure 3a | Figure 2a |
| Bernal et al.[2] | Molnupiravir vs. placebo (twice daily for 5 days) | Outpatients with at least one risk factor for severe illness[b] | Any cause hospitalization or death by day 29 | Nasopharyngeal swabs. Reported as mean change from baseline in $\log_{10}$ copies/mL | 1,3,5,10,15 | Table S6 | Abstract – Results[c] |
| Caraco et al.[3] | Molnupiravir vs. placebo (twice daily for 5 days) | Outpatients, with moderate disease or mild disease and at least one risk factor for severe disease | Any cause hospitalization or death by day 29 | Nasopharyngeal swabs. Reported as mean change from baseline in $\log_{10}$ copies/mL | 1,3,5,10, 15,29 | Figure S2 | Text (Efficacy endpoints) |
| Gottlieb et al. (2021b) [4] | Remdesivir vs. placebo (once daily for 3 days) | Outpatients with at least one risk factor for disease progression | COVID-19 related hospitalization or death from any cause by day 28 | Nasopharyngeal swabs. Reported as mean change from baseline in $\log_{10}$ copies/mL | 1,2,3,7,14[d] | Figure S3 | Table 2 |
| Norton et al.[5] | Casirivimab + Imdevimab vs. placebo (single infusion on day 1) | Outpatients. Overall, 61% of subjects had at least one risk factor for hospitalisation | COVID-19 related hospitalisation or death by day 29 | Nasopharyngeal swabs. Mean change from baseline in $\log_{10}$ copies/mL | 1,3,5,7 | Figure S6A[e] | Table S5[e] |

| | | | | | | | |
|---|---|---|---|---|---|---|---|
| Montgomery et al.[6] | Tixagevimab + Cilgavimab vs. placebo (2 consecutive injections on day 1) | Outpatients. Overall, 90% of subjects were at high risk of progression to severe COVID-19 | Severe COVID-19 (WHO progression score of >=5, which is hospitalization and receiving oxygen), or death by day 29 | Mid turbinate nasal swabs. Mean viral RNA values at each timepoint in $log_{10}$ copies/mL | 1,3,6,15[g] | Figure S1b | Table 2[f] |
| Gottlieb et al. (2021a)[7] | Bamlanivimab (with and without Etesevimab) vs. placebo (single infusion on day 1) | Outpatients. Overall, 67% of subjects had risk factors for severe COVID-19 | COVID-19 related ED visit, hospitalization, or death by day 29 | Nasopharyngeal swabs. Mean change from baseline in $log_{10}$ copies/mL | 1,3,7,11 | Figure 2 | Table 2 |
| Dougan et al. (2021)[8] | Bamlanivimab and Etesevimab vs. placebo (single infusion on day 1) | Outpatients with at least one risk factor for severe disease | COVID-19 related hospitalization, or death from any cause by day 29 | Nasopharyngeal swabs. Mean change from baseline in $log_{10}$ copies/mL | 1,3,5,7,11 | Figure 3 | Results heading of abstract[h] |
| Dougan et al. (2022a)[9] | Bebtelovimab, alone, or with Bamlanivimab and Etesevimab vs. placebo (single infusion on day 1) | Outpatients with no risk factors for severe disease[i] | COVID-19 related hospitalization, or death from any cause by day 29 | Nasopharyngeal swabs. LS mean change from baseline in $log_{10}$ copies/mL | 1,3,5,7,11 | Table 2, Figure 3 | Table 2 |
| Streinu-Cercel et al.[10] | Regdanvimab vs. placebo (single infusion on day 1) | Outpatients. Overall, 71% of subjects were at high risk of progression to severe COVID-19 | COVID-19 related hospitalization or oxygen therapy or death by day 28 | Nasopharyngeal swabs. Mean change from baseline in $log_{10}$ copies/mL | 1,2,3,4,5,6,7,10,14,17,21,28 | Figure S2 | Table 2 |

| | | | | | | | |
|---|---|---|---|---|---|---|---|
| Dougan et al. (2022b)[11] | Bamlanivimab and Etesevimab vs. placebo (single infusion on day 1) | Outpatients with at least one risk factor for severe disease | COVID-19 related hospitalization, or death from any cause by day 29 | Nasopharyngeal swabs. LS mean change from baseline in $\log_{10}$ copies/mL | 1,3,5,7,11 | Figure 3 | Figure 2a |
| Chew et al.[12] | Bamlanivimab vs. placebo (single infusion on day 0) | Outpatients. Overall, 48% of subjects were at high risk of progression to severe COVID-19 | COVID-19 related hospitalization, or death from any cause by day 28 | Nasopharyngeal swabs. Median viral RNA values at each timepoint in $\log_{10}$ copies/mL | 0,3,7,14,21,28 (1,4,8,15,22,29)[j,k] | Table 3 | Table S9 |
| Rossignol et al.[13] | Nitazoxanide vs. placebo (twice daily for 5 days) | Outpatients. Overall, 63% of subjects were at risk of severe COVID-19 per CDC guidelines | COVID-19 related hospitalization or death by day 28 | Nasopharyngeal swabs. Mean change from baseline in $\log_{10}$ copies/mL | 1,4,10[j] | Table 5 | Table 2 |
| Mitja et al. [14] | Hydroxychloroquine (daily for 7 days) vs. usual care | Outpatients. Overall, 53% had "any comorbidity" | COVID-19 related hospitalization or death by day 28 | Nasopharyngeal swabs. Mean change from baseline in $\log_{10}$ copies/mL | 1,3,7 | Table 2 | Table 2 |

[a] Used mITT population which was all subjects treated <= 3 days from symptom onset.

[b] Used final (all randomized) rather than interim results.

[c] We used number of events from abstract, which differs by 1 to numbers shown in legend of Kaplan-Meier curve (Figure 2) due to 1 participant being censored.

[d] Figure S3 shows baseline as day 0, however we have determined this was day 1 as per study protocol section 6.3.

[e] Baseline characteristics and clinical outcome based on the modified full analysis set (mFAS) for groups 1&2 combined, virological outcomes based on mFAS for group 2 only. Virological outcomes for group 1 mFAS are reported separately. [15]

[f] Used full analysis set for clinical outcome (third supportive estimand from table 2).

[g] Supp Figure 1 has baseline labelled as day 0 on chart axis, however we have determined this was day 1 based on data below figure and description of procedures provided.

[h] Clinical outcome numbers differ between primary outcome reported in text and legend of figure 2. We have used primary outcome reported in text.

[i] Only the placebo controlled low risk arms of this trial were used.

[j] Study not used in models analysing virological effects at day 3, 5, and 7 because no virological swabs reported on those days.

[k] This study defined the baseline as day 0, in contrast to the other studies that define this as day 1. We have adjusted the numbering of days in this study to start from day 1 in line with other studies.

**Table S2** Summary of all model results

**Summary of all model results (fold drop)**

| Model description | model estimate ($\beta$)[a] | OR[a] | 95% CI | p-value |
|---|---|---|---|---|
| **Single timepoint models Unadjusted** | | | | |
| Day 3 | -0.62 | 0.54 | 0.38 to 0.74 | 0.0002 |
| Day 5 | -0.61 | 0.54 | 0.41 to 0.72 | <0.0001 |
| Day 7 | -0.091 | 0.91 | 0.74 to 1.13 | 0.39 |
| **Adjusted for treatment type** | | | | |
| Day 3 | -0.61 | 0.54 | 0.39 to 0.76 | 0.0003 |
| Day 5 | -0.59 | 0.56 | 0.41 to 0.74 | <0.0001 |
| Day 7 | -0.079 | 0.92 | 0.75 to 1.14 | 0.46 |
| **Adjusted for variant** | | | | |
| Day 3 | -0.62 | 0.54 | 0.39 to 0.75 | 0.0002 |
| Day 5 | -0.61 | 0.55 | 0.41 to 0.72 | <0.0001 |
| Day 7 | -0.097 | 0.91 | 0.74 to 1.12 | 0.36 |
| **Composite models** | | | | |
| Maximum virological effect model | -0.25 | 0.78 | 0.65 to 0.92 | 0.0038 |
| Average virological effect model | -0.27 | 0.76 | 0.61 to 0.96 | 0.019 |
| Clearance rate model | -0.35 | 0.71 | 0.55 to 0.90 | 0.0052 |

[a] $\beta$ and OR are for a two-fold increase in virological treatment effect in treated subjects in all models except the viral clearance model, where we report the estimated change in efficacy for an increase in clearance rate of 0.1 $\log_{10}$ copies/mL per day.

**Table S3** Sensitivity analysis for high RoB studies

| Model description | model estimate ($\beta$)[a] | OR[a] | 95% CI | p-value |
|---|---|---|---|---|
| **Single timepoint models – adjusting for RoB** | | | | |
| Day 3 | -0.62 | 0.54 | 0.38 to 0.74 | 0.0002 |
| Day 5 | -0.61 | 0.54 | 0.41 to 0.72 | <0.0001 |
| Day 7 | -0.15 | 0.86 | 0.70 to 1.06 | 0.15 |

[a] $\beta$ and OR are for a two-fold increase in virological treatment effect in treated subjects.

**Table S4** methods used to convert reported measures of virological uncertainty to 95% CIs for virological treatment effect.

| Data conversion index | Description of what was reported/extracted from original study | Conversion used to get 95% Confidence Interval (CI) | Papers |
|---|---|---|---|
| C0 | 95% CI of difference reported | Use reported 95% CI | Gottlieb et al. (2021a) |
| C1 | standard error of mean difference between groups | 1.96*se | Hammond et al. |
| C2 | Standard error of mean change from baseline separately for each group | $se(diff) = \sqrt{se_1^2 + se_2^2}$ Then proceed as in (C1) | Streinu-Cercel et al. Dougan et al. (2022b) Dougan et al. (2022a) Rossignol et al. Mitja et al. |
| C3 | standard deviation of mean change from baseline separately for each group (and *n* participants) | $se = sd/\sqrt{n}$ Then proceed as in (C2) | Bernal et al. |
| C4 | 95% CI of change from baseline separately for each group | $se = \frac{0.5 \times CI_{width}}{1.96}$ Then proceed as in (C2) | Gottlieb et al. (2021b) Montgomery et al. Dougan et al. (2021) |
| C5 | No standard deviation, standard error or confidence intervals provided | 95% CI in treatment effect not calculated | Norton et al. Caraco et al. Chew et al. |

**Figure S1** Risk of Bias assessment summaries for (A) clinical outcomes and (B) virological outcomes.

A

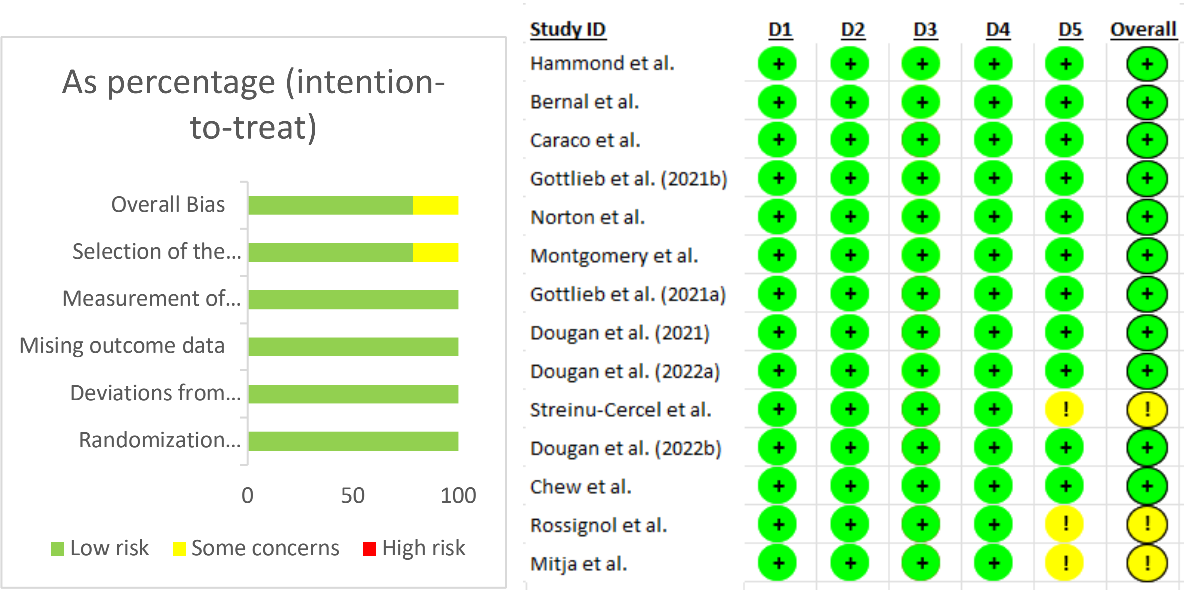


B

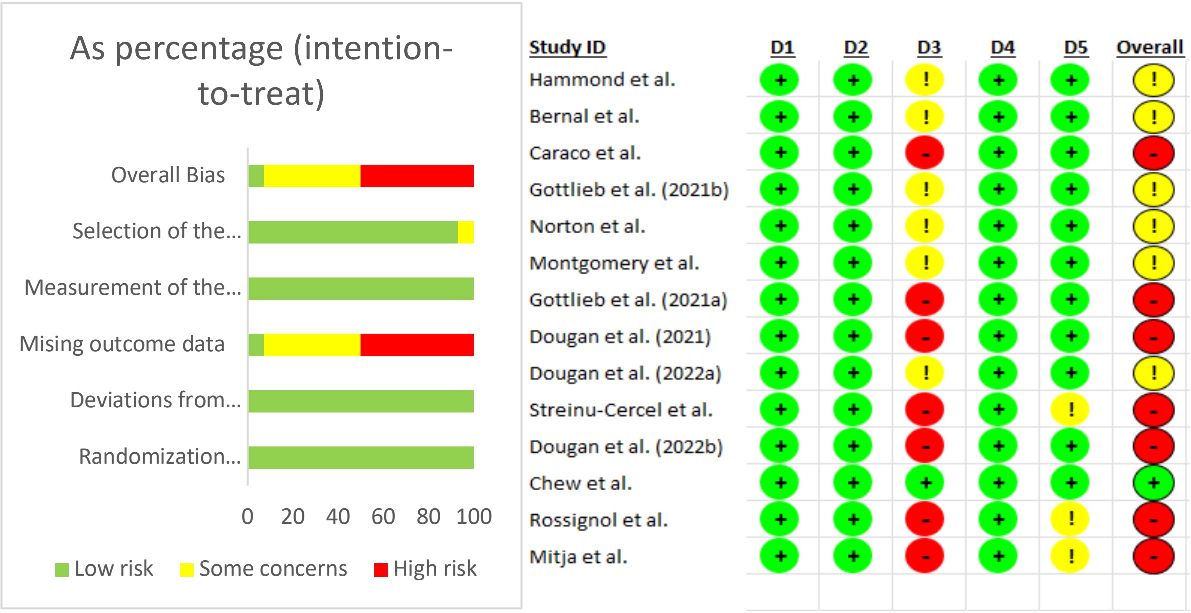


**Domains**
D1: Bias arising from the randomization process
D2: Bias due to deviations from intended intervention
D3: Bias due to missing data
D4: Bias in measurement of the outcome
D5: Bias in selection of the reported result

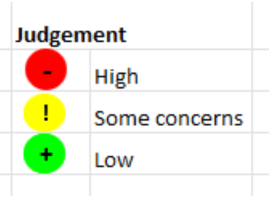

**Figure S2** Available timepoints for viral load data

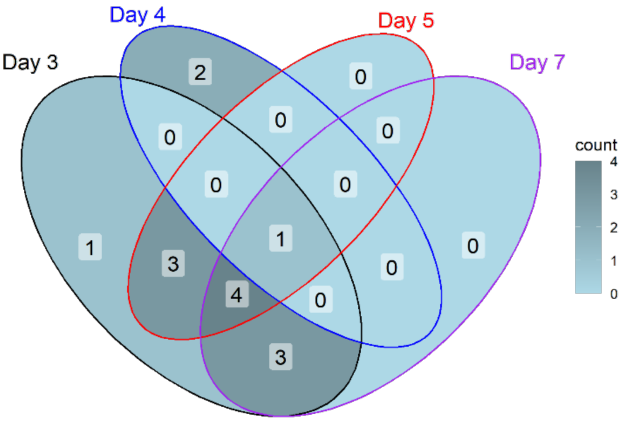

**Figure S3** Correlation between virological effects of treatment observed at different timepoints.

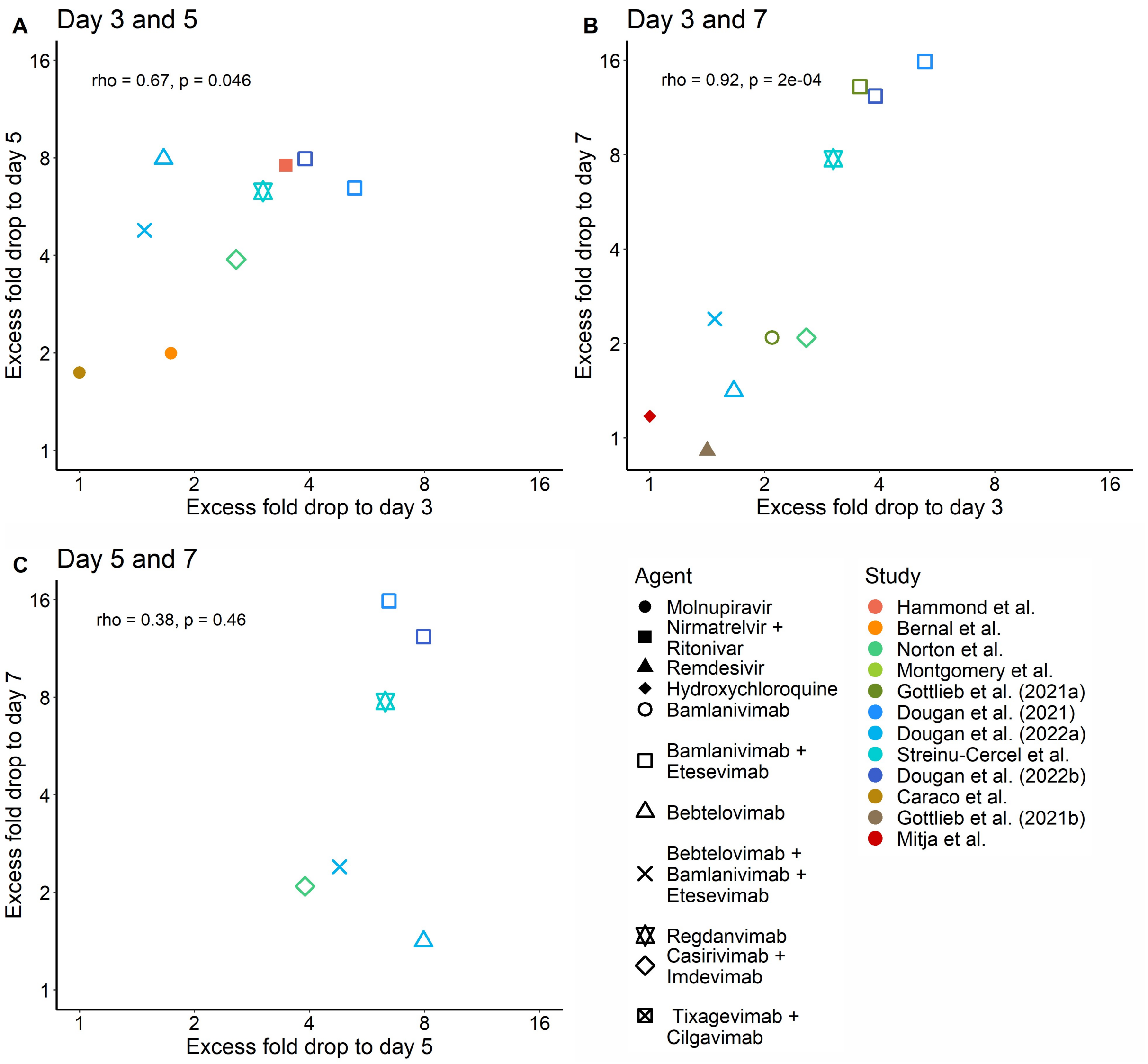


**Figure S3** Relationship between the virological effects of treatment (fold-drop in viral load in treated vs controls) based on different viral load measurement times. Relationship is shown for all studies with viral load data available at both timepoints for (A) days 3 and 5 (n=9), (B) days 3 and 7 (n=10) and (C) days 5 and 7 (n=6). Pearson correlation statistics are shown on each panel.

**Figure S4** Composite models of virological treatment effect

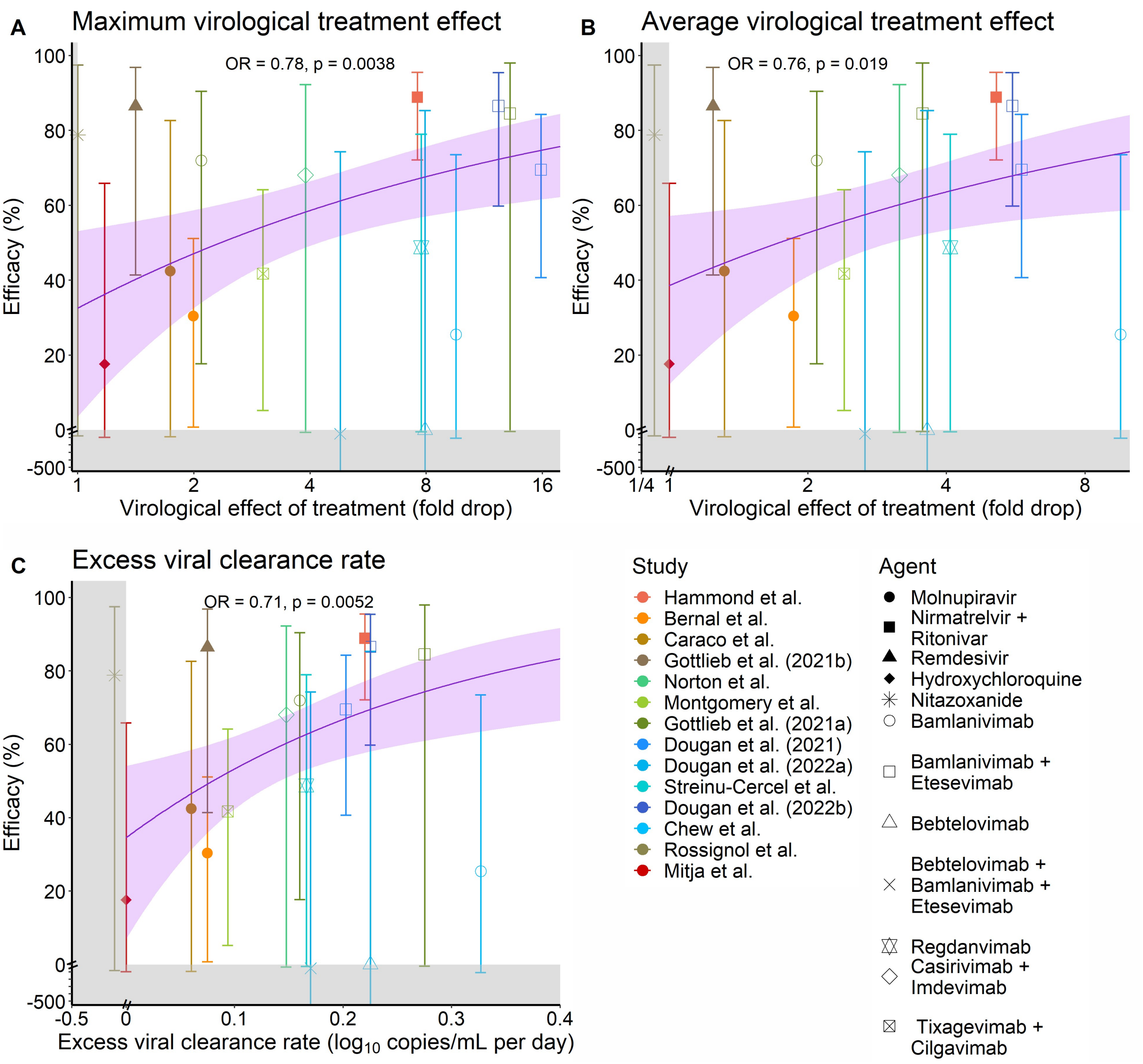


**Figure S4** Relationship between virological treatment effect and clinical efficacy predicted by various composite measures of the virological treatment effect, which were then used as predictors in a GLMM (in the same way as with the single timepoint models in Fig 4). Composite measures used were (A) the maximum observed virological treatment effect up to and including day 7, (B) the average virological treatment effect using all available data up to and including day 6; and (C) the excess viral clearance rate estimated by OLS for each study using all available virological data up to and including day 6. Error bars are the 95% confidence intervals. The solid line indicates the fitted model, and the shaded region around this line indicates the 95% confidence region. Grey shading indicates an axis break point.

**Figure S5** Predicting the virological treatment effects required to have confidence in a given level of clinical protection.

For example, if the primary outcome of a study is virological treatment effect on day 3, then a 2.3-fold larger drop in viral load is required in the treated arm compared with control arm (black dotted line) in order to have 95% confidence in greater than or equal to 50% protection. e.g. the lower bound of the 95% confidence interval shows a lower confidence interval of 50% clinical efficacy (bottom of shaded region in Figure S5A). This can be interpreted as saying that if we believe the true virological treatment effect at day 3 is at least an extra 2.3-fold drop (1.2 Ct), then we have 95% confidence of achieving a clinical efficacy of at least 50%, and our best estimate of clinical efficacy is 60% (95% CI 50% to 67%). Alternatively, if the desired efficacy is at least 70%, then a virological treatment effect of an extra 5.7-fold drop (2.5 Ct) at day 3 (red dotted line) gives 95% confidence of a treatment efficacy greater than or equal to 70% (and an expected clinical efficacy of 82% (95% CI 70% to 90%)). Similar analysis can also be provided based on the virological treatment effects at day 5 post-treatment (Figure S5B).

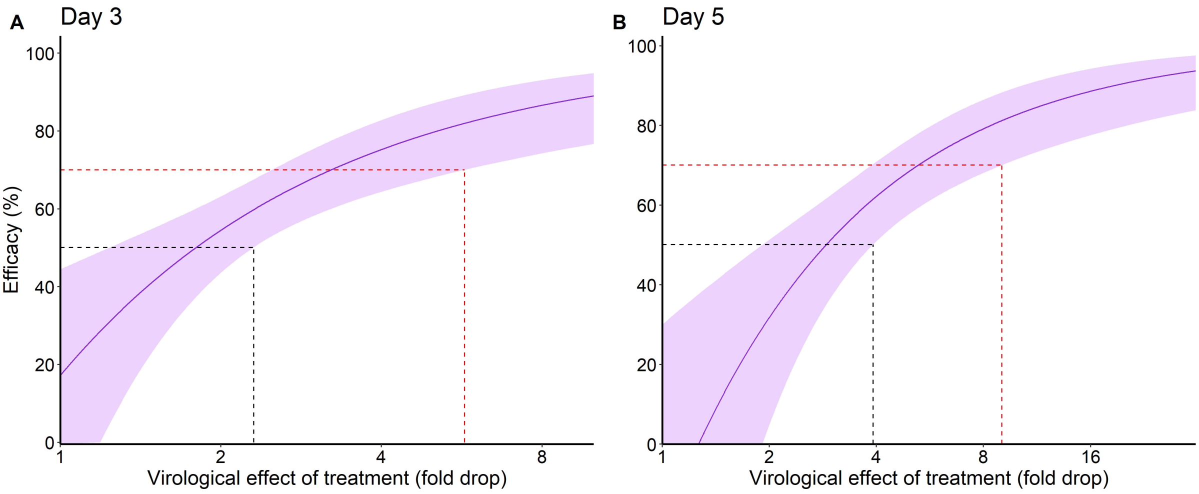


**Figure S5** Estimating efficacy using models of virological effect at (A) day 3 and (B) day 5.

### References

1. Hammond J, Leister-Tebbe H, Gardner A, et al. Oral Nirmatrelvir for High-Risk, Nonhospitalized Adults with Covid-19. *N Engl J Med* 2022; **386**(15): 1397-408.
2. Jayk Bernal A, Gomes da Silva MM, Musungaie DB, et al. Molnupiravir for Oral Treatment of Covid-19 in Nonhospitalized Patients. *New England Journal of Medicine* 2021; **386**(6): 509-20.
3. Caraco Y, Crofoot GE, Moncada PA, et al. Phase 2/3 Trial of Molnupiravir for Treatment of Covid-19 in Nonhospitalized Adults. *NEJM Evidence* 2022; **1**(2): EVIDoa2100043.
4. Gottlieb RL, Vaca CE, Paredes R, et al. Early Remdesivir to Prevent Progression to Severe Covid-19 in Outpatients. *New England Journal of Medicine* 2021; **386**(4): 305-15.
5. Norton T, Ali S, Sivapalasingam S, et al. REGEN-COV Antibody Combination in Outpatients With COVID-19 – Phase 1/2 Results. *medRxiv* 2022: 2021.06.09.21257915.
6. Montgomery H, Hobbs FDR, Padilla F, et al. Efficacy and safety of intramuscular administration of tixagevimab-cilgavimab for early outpatient treatment of COVID-19 (TACKLE): a phase 3, randomised, double-blind, placebo-controlled trial. *Lancet Respir Med* 2022.
7. Gottlieb RL, Nirula A, Chen P, et al. Effect of Bamlanivimab as Monotherapy or in Combination With Etesevimab on Viral Load in Patients With Mild to Moderate COVID-19: A Randomized Clinical Trial. *JAMA* 2021; **325**(7): 632-44.
8. Dougan M, Nirula A, Azizad M, et al. Bamlanivimab plus Etesevimab in Mild or Moderate Covid-19. *New England Journal of Medicine* 2021; **385**(15): 1382-92.
9. Dougan M, Azizad M, Chen P, et al. Bebtelovimab, alone or together with bamlanivimab and etesevimab, as a broadly neutralizing monoclonal antibody treatment for mild to moderate, ambulatory COVID-19. *medRxiv* 2022: 2022.03.10.22272100.
10. Streinu-Cercel A, Săndulescu O, Preotescu LL, et al. Efficacy and Safety of Regdanvimab (CT-P59): A Phase 2/3 Randomized, Double-Blind, Placebo-Controlled Trial in Outpatients With Mild-to-Moderate Coronavirus Disease 2019. *Open Forum Infect Dis* 2022; **9**(4): ofac053.
11. Dougan M, Azizad M, Mocherla B, et al. A Randomized, Placebo-Controlled Clinical Trial of Bamlanivimab and Etesevimab Together in High-Risk Ambulatory Patients With COVID-19 and Validation of the Prognostic Value of Persistently High Viral Load. *Clin Infect Dis* 2022; **75**(1): e440-e9.
12. Chew KW, Moser C, Daar ES, et al. Antiviral and clinical activity of bamlanivimab in a randomized trial of non-hospitalized adults with COVID-19. *Nature Communications* 2022; **13**(1): 4931.
13. Rossignol J-F, Matthew CB, Oaks JB, et al. Early treatment with nitazoxanide prevents worsening of mild and moderate COVID-19 and subsequent hospitalization. *medRxiv* 2021: 2021.04.19.21255441.
14. Mitjà O, Corbacho-Monné M, Ubals M, et al. Hydroxychloroquine for Early Treatment of Adults With Mild Coronavirus Disease 2019: A Randomized, Controlled Trial. *Clin Infect Dis* 2021; **73**(11): e4073-e81.
15. Weinreich DM, Sivapalasingam S, Norton T, et al. REGN-COV2, a Neutralizing Antibody Cocktail, in Outpatients with Covid-19. *New England Journal of Medicine* 2020; **384**(3): 238-51.
